# Development of a Tremor Detection Algorithm for use in an Academic Movement Disorders Center

**DOI:** 10.1101/2024.03.13.24304101

**Authors:** Mark Saad, Sofia Hefner, Suzann Donovan, Doug Bernhard, Richa Tripathi, Stewart A. Factor, Jeanne Powell, Hyeokhyen Kwon, Reza Sameni, Christine D. Esper, J. Lucas McKay

## Abstract

Tremor, defined as an “involuntary, rhythmic, oscillatory movement of a body part,” is a key feature of many neurological conditions, but is still clinically assessed by visual observation. Methodologies for objectively quantifying tremor are promising but remain non-standardized across centers. Our center performs full-body behavioral testing with 3D motion capture for clinical and research purposes for patients with Parkinson’s disease, essential tremor, and other conditions. The objective of this study was to assess the ability of several candidate processing pipelines to identify the presence or absence of tremor in kinematic data from movement disorders patients compared to expert ratings from movement disorders specialists. We curated a database of 2,272 separate kinematic data recordings from our center, each of which was contemporaneously annotated as tremor present or absent by a clinical provider. We compared the ability of six separate processing pipelines to recreate clinician ratings based on F1 score, in addition to accuracy, precision, and recall. We found generally comparable performance across algorithms. The average *F*1 score was 0.84 *±* 0.02 (Mean *±*SD; range 0.81 *−* 0.87), with all *F*1 confidence intervals overlapping. The highest performing algorithm (cross-validated *F*1 = 0.87) was a hybrid that used engineered features adapted from an algorithm in longstanding clinical use with a modern Support Vector Machine classifier. Taken together, our results suggest the potential to update legacy clinical decision support systems to incorporate modern machine learning classifiers in order to create better performing tools.

## 1. Introduction

Tremor, defined as an “involuntary, rhythmic, oscillatory movement of a body part,” is a key feature of many neurological conditions, and has been called the most frequent human movement disorder [1]. In Parkinson’s disease (PD), the second most common neurodegenerative disorder worldwide [2], tremor that appears while at rest (often a “pill-rolling” tremor of the thumb and forefinger) is considered a characteristic sign [3]. However, tremor is a feature of many neurological conditions [3], and can also result from various causes such as trauma or side effects of medications [4]. Furthermore, some oscillatory movements occur that are not tremor; myoclonus and dystonia can produce involuntary, jerking movements that may be rhythmic, but that are not considered to be tremor [5].

Tremor disorders currently require expert diagnosis based on skilled observation using standardized clinical scales, with quantitative measurements approximated by eye and with no automated clinical decision support. Clinicians characterize the features of the tremor, including body distribution, provocative factors, frequency, and gross amplitude and aggregate this information with other medical testing results to identify underlying causes and to evaluate potential treatment plans[2]. In PD and Essential Tremor (ET), overall tremor severity is measured using standardized clinical scales like the Movement Disorder Society-Unified Parkinson’s Disease Rating Scale Part III (MDS-UPDRS-III) [6], the Fahn-Tolosa-Marin Tremor Rating Scale (FTM) [5] and The Essential Tremor Rating Scale (TETRAS) [7]. These give general guidelines for tremor amplitude assessment by eye, but are not intended to be used with actual measurements (e.g., with calipers or an anthropometer).

Recent progress in human activity recognition and edge computing suggests significant potential for automated clinical decision support tools in tremor measurement. Despite this, technologies for identifying tremor have progressed towards standardization and clinical uptake very slowly [1]. In the research domain, various technologies measure human motion, including body-worn sensors [3,7,8], 3D motion capture [9,10], and – most recently – pose recognition from monocular video [11–13]. Digitizing tablets are often used for assessing tremor during tasks like spiral drawing [14,15] and for discriminating tremor from bradykinesia during finger tapping [16]. In fact, recognition of the potential for spectral analysis in assessing tremor dates back to the mid-1960s [17], and differences in tremor frequencies across disorders have been acknowledged for over two decades [3]. Substantial domain knowledge (and in some cases, cultural) gaps between clinicians and engineers further hampers widespread adoption. This is in contrast to fields like cardiology, where automated clinical decision support systems thrive due to large public datasets enabling annual improvements in anomaly detection [18,19].

In our center, we perform comprehensive behavioral testing using 3D kinematic motion capture to objectively evaluate abnormal movements in patients with PD, ET, and other conditions [20]. Indications for this procedure include diagnosis adjudication and evaluation for functional neurosurgery, among others. Our behavioral testing paradigm involves multiple standardized upper limb tasks designed to elicit tremor under provoking conditions of rest, posture, and action. Since 2014, we have performed >1500 behavioral tests, using analysis pipelines that were developed organically based on clinician domain knowledge without formal evaluation. A challenge encountered in evaluating tremor analysis algorithms is imprecision in the “ground truth” criteria for tremor presence outlined in clinical scales [5–7]. For instance, the MDS-UPDRS-III criterion that “tremor is present but less than 1 cm in amplitude” (corresponding to a score of 1) is clear for a human rater but poses ambiguity for a machine. Questions arise in implementation, such as along which biomechanical axis or axes the amplitude should be measured and what size tremor meets the threshold for being considered “present.”

The objective of this study is to compare tremor identification algorithms from our clinic for identifying tremors in 3D kinematic data of movement disorder patients. Ground truth labels, recorded in contemporaneous notes by clinicians, are straightforward: tremor present or absent. The main goal is to identify the most accurate algorithm for detecting tremor presence or absence in individual body parts during testing sessions.

## 2. Materials and Methods

### 2.1. Data sources

We compared algorithm performance using a database of 2,272 recordings made during standard clinical exams of N = 50 arbitrarily selected clinic patients. Aspects of the testing paradigm have been described previously [9,20,21]; more detail is provided below. In 42 patients (84%) the primary diagnosis was either Parkinson’s disease or essential tremor. Detailed demographic and clinical characteristics are shown in Table 1.

**Table 1.** Clinical and demographic characteristics of the study sample.

| Variable | Value |
| --- | --- |
| N | 50 |
| Sex |  |
| Male | 32 (64%) |
| Female | 18 (36%) |
| Age, years |  |
| Mean (SD) | 65 (12) |
| Range | 36 - 83 |
| Primary Diagnosis |  |
| Parkinson's disease | 29 (58%) |
| Essential tremor | 13 (26%) |
| Dystonia | 6 (12%) |
| Essential tremor + Parkinson's disease | 1 (2%) |
| Unspecified tremor | 1 (2%) |

### 2.2. Behavioral testing paradigm

Behavioral testing was captured through 3D optical motion capture with 60 reflective markers on standardized bony landmarks during a 1-hour clinical assessment in our facility (Figure 1). Assessments were billed under Current Procedural Terminology (CPT [22]) codes 96000 and 96004. All patients with Parkinson’s disease were asked to hold their antiparkinsonian medications for at least 12 hours prior to the study visit (the practically-defined OFF state [23]). At the time of testing, the average time since the last medication dose was 13±5 hours. Tasks included goal-directed upper limb movements, static postures, and walking, and were designed to provoke various tremors [2]. For instance, seated finger-to-nose pointing with the right arm (coded *sit-point-right* or *sit-point-1* in data files) aimed to elicit action-provoked tremor in the right upper extremity and rest or postural tremor in the legs, left upper extremity, torso, head, and neck (Table B.2). On average, kinematic data recordings were 27 *±* 9 seconds long and ranged from 3 to 92 seconds.

**Figure 1.**
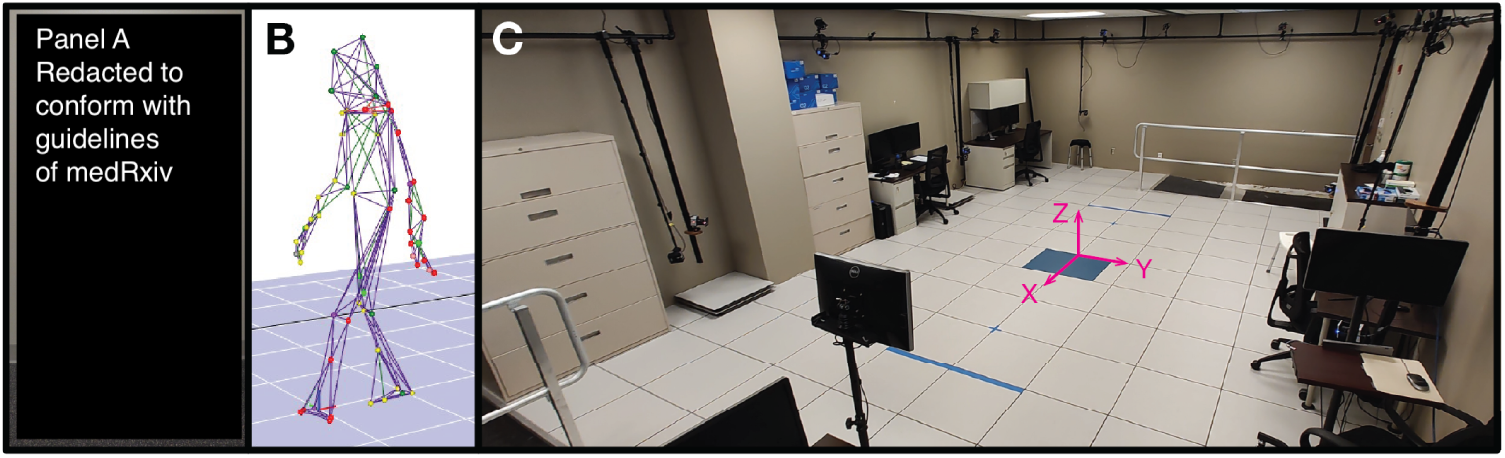
Clinical motion capture facility. Our center uses a custom set of 60 retroreflective kinematic markers for most cases. Markers on the hands (arrows) enable tremor measurement. From top to bottom, the markers highlighted are R.Wrist, R.Thumb.M3, and R.Finger3.M3 (A). After data collection, analysis is performed using a deidentified wire frame or representation of the individual, preserving privacy (B). Our 650 square feet center is used for both clinical and research applications (C). The origin of the kinematic coordinate system is superimposed.

**Table 2.** Comparison of tremor identification algorithms. All algorithms operate on spectral features of kinematic data.

| Algorithm | Kinematic Data | Features | Aggregation | Classifier |
| --- | --- | --- | --- | --- |
| A1r<br>A1s | Velocity | Engineered | Winner-take-all | Rule-Based<br>SVM |
| A2r<br>A2s | Position | Engineered | Average | Rule-Based<br>SVM |
| B1 | Position | Data-driven | Average | SVM |
| B2 | Position | Data-driven | Average | XGBoost |

### 2.3. Kinematic data recording, processing, and export

Data were captured using a 3D motion capture system (Motion Analysis Corporation, Rohnert Park, CA, USA) with 10 cameras recording at 120 Hz. Following testing completion, clinic staff manually postprocessed kinematic data using standard interpolation features in Motion Analysis Cortex software for quality control. Occasional low-pass or similar filters were applied on an as-needed basis to address noise in individual markers, but no consistent additional filtering occurred. Each recording’s kinematic data was exported into standard *\*.trc* tabular format. A typical *.trc* file for a 30-second recording at 120 Hz comprises 3600 rows (30 seconds × 120 Hz) and 180 columns (60 markers × 3 axes) of kinematic data. Due to changes in marker labels and occasional missing data, each *.trc* file was divided into separate *.csv* files for each body extremity in the accompanying dataset. These files are compatible with standard Python, R, Matlab, or similar software libraries. Summaries of the contents of example files are provided in Table B.1.

### 2.4. Annotations

Annotations were taken contemporaneously during the exam for the clinicians’ own use while preparing exam notes. Because tremor is intermittent in nature and typically does not appear across more than a few isolated body regions, annotations typically included separate entries for particular body parts during each particular recording. For example, the annotation “Left hand: present, F3 and thumb” was used to indicate that tremor was present on the third finger (F3) and thumb of a particular trial. Therefore, the annotations were converted by the study team into separate annotations for each body extremity during each recording. For example, “mild bilateral rest hand tremor” was converted into the annotation “tremor present” for each of the left and right hands. As the presence or absence of tremor in other body extremities was ambiguous in this case, no annotations were provided for other body extremities. In cases where the absence of tremor was described in the original notes (“this gentleman does not have tremor”) tremor was labeled as “tremor absent” for all extremities. Cases in which dyskinesia or dystonic posturing was present were labeled as “tremor absent.”

### 2.5. Spectral composition of kinematic data

All algorithms used initial preprocessing to isolate spectral (or “frequency-domain”) features of recorded data based on the substantial amount of established research in this area. Tremor frequencies vary from 0.5 Hz for palatal tremor to 18 Hz for primary orthostatic tremor, [2] with the majority of parkinsonian and essential tremor typically occurring between 4-12 Hz. [3] Volitional movements during behavioral testing typically occur primarily at lower frequencies, while higher frequency ranges are prone to artifacts related to aliasing or electrical or other noise. For this reason, tremor data are typically processed by band-pass filtering. Typical ranges include 1 to 16, 0.5 to 15, or 2 to 30 Hz [1]. All of the tremor detection algorithms examined employed some initial band-pass or other filtering, described below.

### 2.6. Algorithms

Identifying tremor is a process that uses the rich information embedded within motion data from kinematic markers on each extremity during to determine whether tremor is present or absent in a particular testing session. Although this use case is unique, like many general machine learning problems, this process can be broken down into two basic steps. The first step is *feature engineering*: extracting information (“features“) from raw kinematic marker data. The second step is *classifier development*: creating a classifier based on the extracted features that determines whether tremor is present or absent. During classifier development, in particular, it is important to perform some hyperparameter optimization to identify the optimal operating point for a given algorithm.

In this study, we compare six algorithms for identifying tremor (Table 2). The first two (A1r and A2r) were developed organically over several years based on clinical expertise and signal processing heuristics. As implemented in our clinic, both algorithms derive engineered spectral features from the kinematic data which are then input into simple rule-based classifiers to determine whether tremor is present or absent. While these algorithms were developed iteratively over several years with access to the clinical dataset, no comprehensive hyperparameter tuning was performed, potentially leading to suboptimal parameter settings.

To create a more fair comparison with the modern machine learning algorithms (B1 and B2), we also examined the performance of these algorithms when the features identified by each (summarized in Table 3) were used as inputs to a well-established machine learning model (Support Vector Machines, SVMs [24]) trained and evaluated with 5-fold cross-validation. To distinguish these algorithms from the related algorithms with rule-based classifiers, these implementations are referred to as A1s and A2s.

**Table 3.**
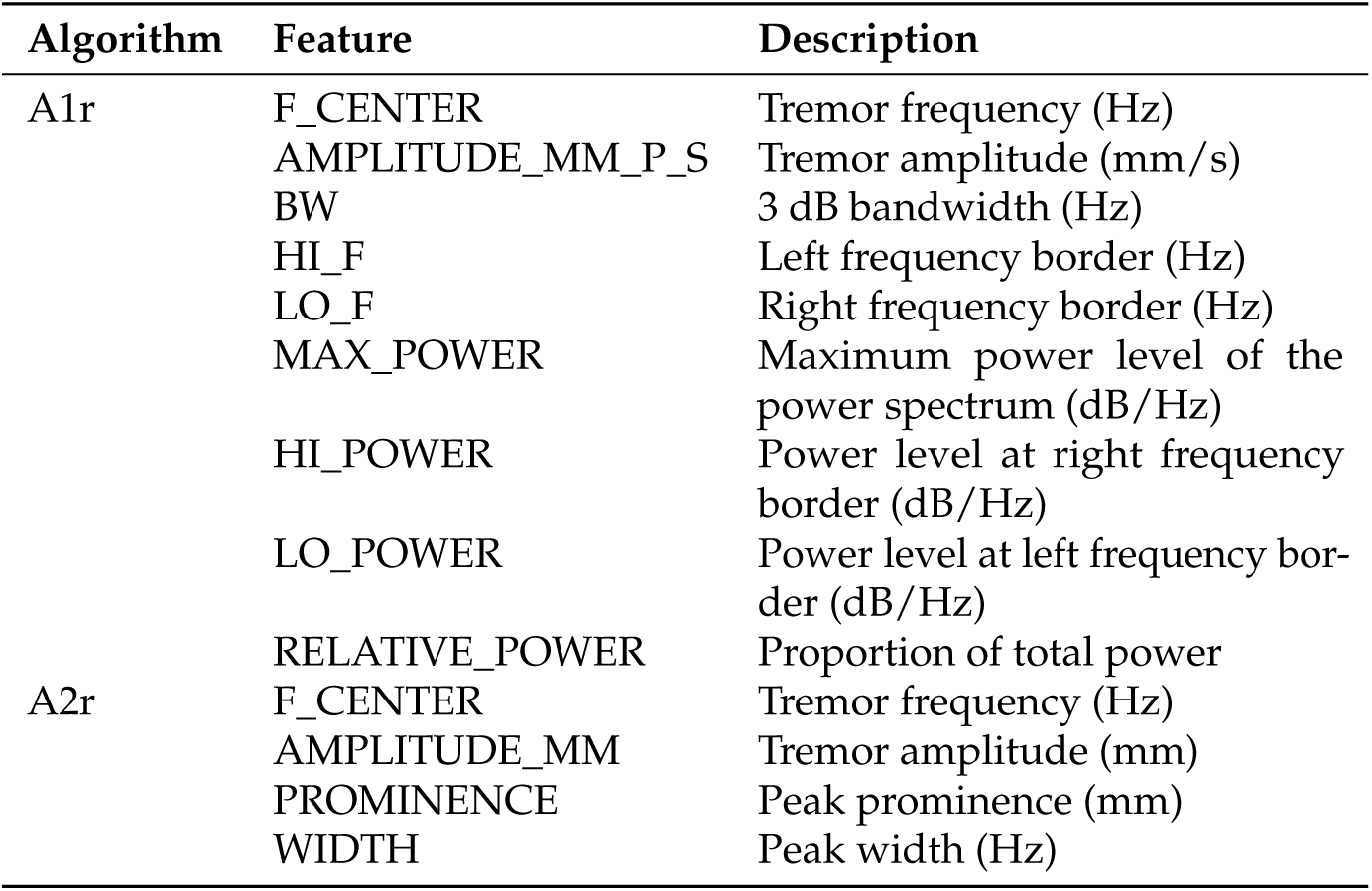
Spectral features calculated by clinical algorithms A12r/s and A2r/s.

The final two algorithms (B1 and B2) were developed specifically for this study based on standard modern machine learning best practices. Both B1 and B2 use basic preprocessing and spectral features together with well-established machine learning models to identify optimal operating points. The details of each algorithm are described below.

#### 2.6.1. Velocity Spectral Peak Detection (Algorithm A1r)

The oldest algorithm in use in our center was developed iteratively between 2014 and 2020. The key feature of this algorithm is that it performs numerical differentiation on kinematic data prior to feature identification in the frequency domain. It uses a winner-take-all approach to aggregate tremor features across kinematic markers on a given extremity (described below). An example of tremor identification using Algorithm A1r is presented in Figure 2. This algorithm was implemented in Matlab (Version R2022b; The Mathworks, Natick, MA, USA).

**Figure 2.**
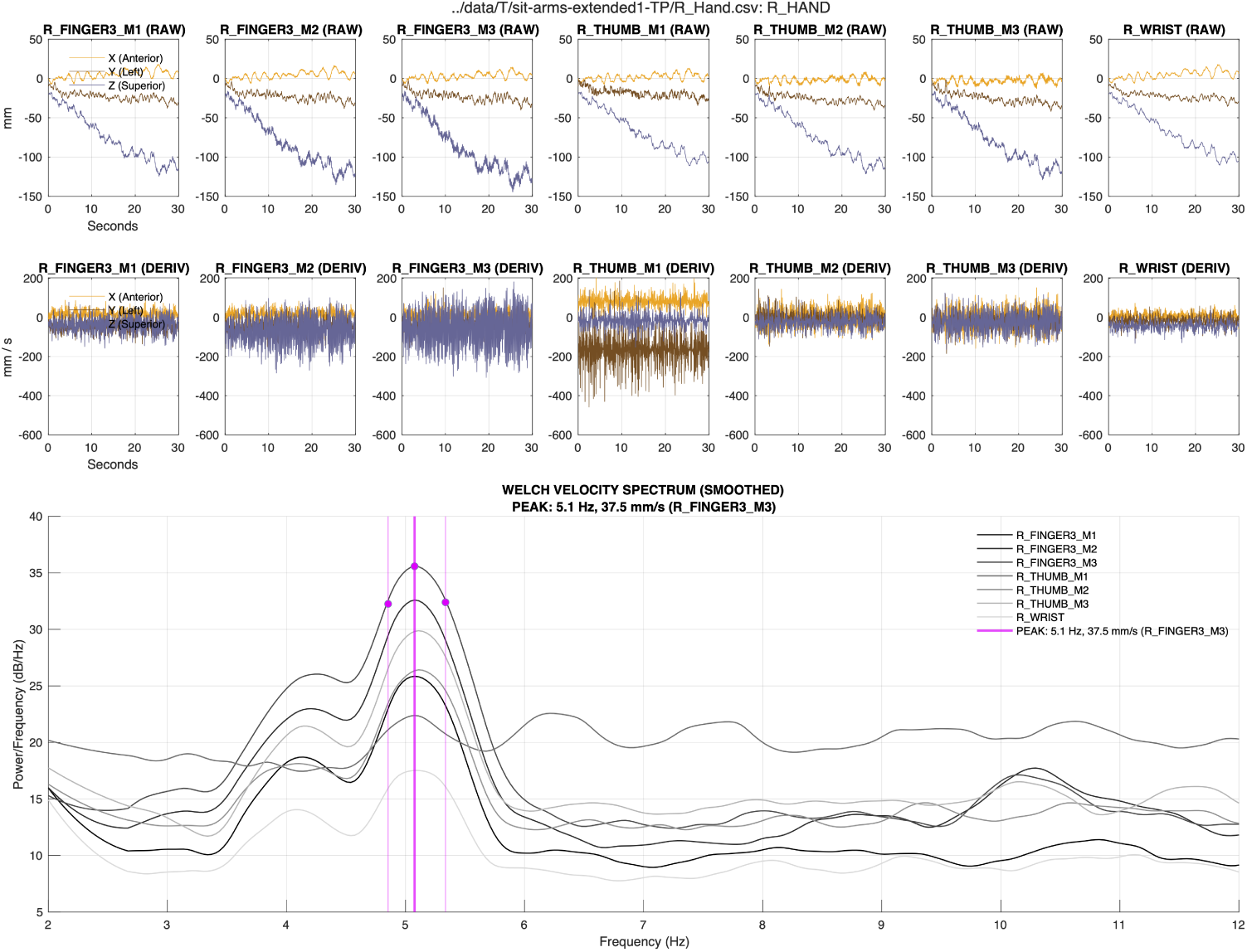
Example of tremor identification with algorithm A1r. Algorithm A1r operates on each kinematic marker on a given extremity, and estimates the central frequency (Hz) and spectral power density (db/Hz) of the highest-amplitude tremor observed across markers.

##### Feature extraction

Raw kinematic displacement data for each marker of a given extremity are zero-phase lowpass filtered (20 Hz), centered, and passed through a Savitzky-Golay derivative filter to obtain smooth velocity estimates in each of the x, y, and z dimensions. The power spectral densities (PSDs) of the velocity components for each marker are obtained using Welch’s method and combined using the Euclidean norm. The combined PSD of each marker is then converted to log scale, smoothed using a Savitzky-Golay filter, and converted back to a linear scale for spectral analysis. Spectral features are summarized in Table 3. More details on feature calculation are available in the documentation for powerbw.m.

##### Rule-based classification

To detect a peak that would indicate the presence of tremor, the peak power and the corresponding center frequency were first detected for each kinematic marker using functionality integrated in the Matlab function powerbw.m. A significant peak should be narrow and symmetric about the center frequency, so any peak with a bandwidth greater than 2 Hz or nonsymmetric power to the left and right of the peak would cast doubt on the presence of a physiological tremor. Indicators of bandwidth and symmetry are derived using powerbw.m and subjected to threshold rules to determine the presence or absence of tremor. Peaks with center frequencies above 10 Hz would also be deemed unreasonable; therefore, central frequencies above 10 Hz are also interpreted as tremor absence. To aggregate features across markers of a given extremity, the algorithm proceeds to detect a tremor on each marker independently. The tremor features for the marker with the largest tremor amplitude on a given recording are used as representative of the entire extremity.

#### 2.6.2. Amplitude Spectral Peak Detection (Algorithm A2r)

The amplitude spectral peak detection was established in our center primarily to provide tremor identification in the amplitude, rather than velocity domain, in order to enable direct comparison with clinical magnitude cutoffs. The key feature of this algorithm is that it converts all kinematic data from kinematic markers on a given extremity to the frequency domain prior to aggregation with a max procedure. Therefore, the spectral features identified for a given extremity reflect a combination of kinematic markers, rather those of a single dominant markers. An example of tremor identification using Algorithm A2r is presented in Figure 3. This algorithm was implemented in Matlab (Version R2022b; The Mathworks, Natick, MA, USA).

**Figure 3.**
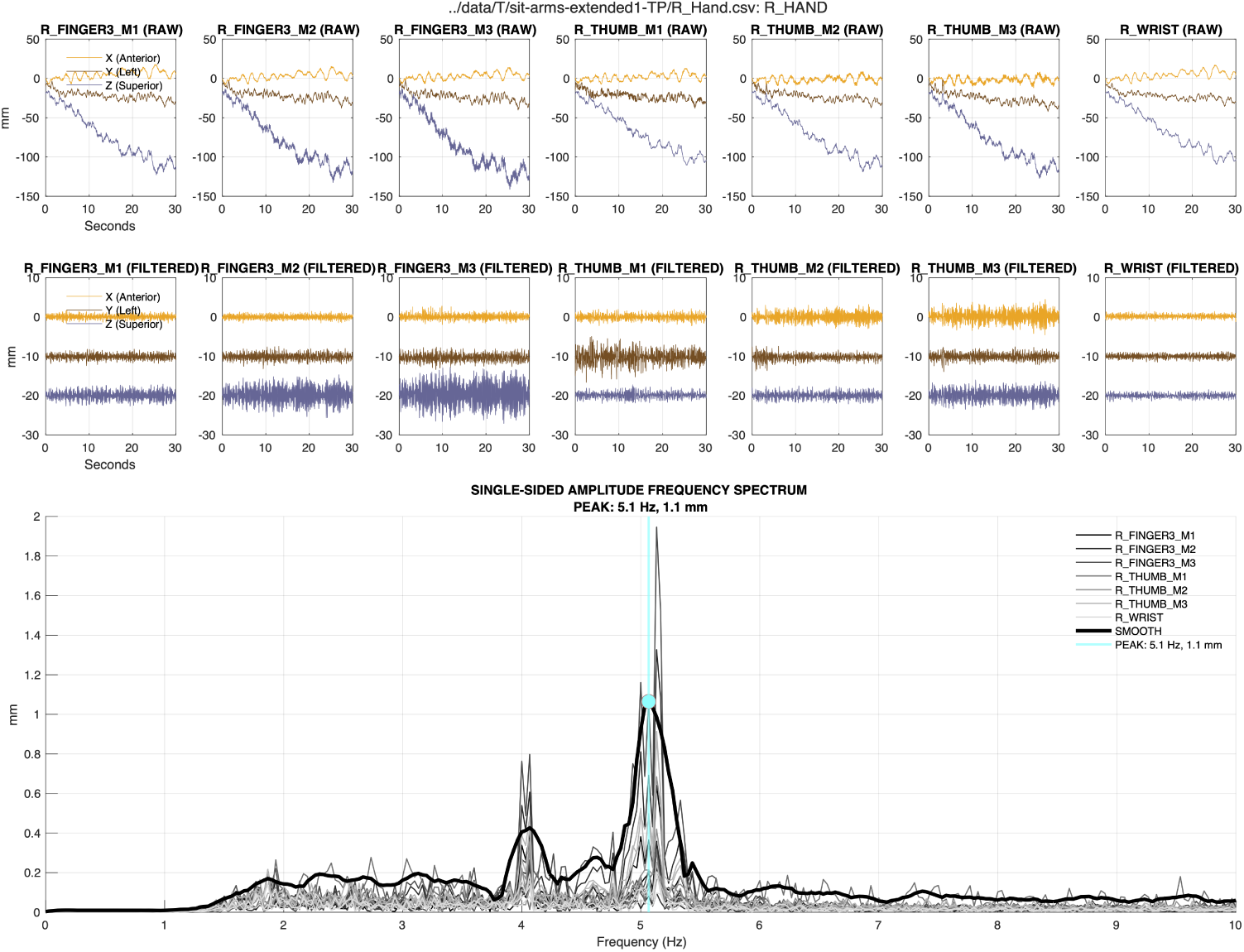
Example of tremor identification with algorithm A2r. Algorithm A2r operates simultaneously on all kinematic markers on a given extremity, and estimates the central frequency (Hz) and amplitude (mm) of the highest-amplitude tremor present.

##### Feature extraction

Raw kinematic data of all markers on a given extremity are high-pass filtered with a 4th order Butterworth filter with corner frequency 2 Hz using filtfilt.m in Matlab. The two-sided frequency spectrum is calculated using the fast Fourier transform and converted into the single-sided frequency spectrum of each axis of each kinematic marker. The single-sided frequency spectra of each x, y, z component of all markers on each extremity are combined using a max procedure to create an aggregate spectrum for the extremity that represents the most severe tremor at each frequency. The aggregate spectrum is subsequently smoothed with a Savitsky-Golay 3rd-order polynomial smoothing filter.’Frequency peaks in the smoothed spectrum are then identified with the heuristic-based findpeaks.m method in Matlab software using default arguments.

##### Rule-based classification

Classification proceeds in two steps. First, the central frequency of the dominant frequency peak identified by findpeaks.m is compared to maximum and minimum threshold values. Peaks with central frequency < 3.5 Hz or > 10 Hz are considered non-physiological and are discarded. If these conditions are met, the amplitude of the peak is compared to the a simple threshold value (0.1 mm) to determine tremor presence. This threshold value was determined over trial and error.

#### 2.6.3. Support Vector Machines with Engineered Spectral Features (Algorithms A1s and A2s)

We also examined the performance of Algorithms A1r and A2r when the final classification steps were altered from the heuristic rule-based implementations to Support Vector Machines (SVMs). Support vector machines (SVM) are a widely recognized approach for classification tasks [25]. An SVM is a supervised machine learning algorithm that works by identifying an optimal hyperplane in an augmented feature domain that separates observations into distinct classes. In this case, observations that fall on one or other side of the hyperplane are classified as tremor present or absent. Importantly, the feature domain can be augmented with features derived via nonlinear functions (here, radial basis functions) in order to accommodate linearly-non separable classes in the original data. Here, we extracted the spectral features identified by each algorithm (summarized in Table 3) and used them as inputs to two separate SVMs with 5-fold cross-validation and radial basis function kernels.

#### 2.6.4. Modern Classifiers (Algorithms B1 and B2)

The final two algorithms (B1 and B2) were developed specifically for this study. They use basic preprocessing and spectral features together with well-established machine learning models to identify tremor in kinematic data.

##### Feature extraction

In order to decouple tremulous movements from slow body movements due to subjects’ displacements and non-tremulous activities, the vector position of each kinematic marker on a given extremity is initially calculated as a measure of its instantaneous distance from the origin of the kinematic reference frame. This is done by calculating the Euclidean norm of the x, y, and z coordinates at all time instants, resulting in a single signal per sensor, as a function of time. The resulting signals are bandpass filtered between 1 Hz and 20 Hz with a linear-phase finite impulse response (FIR) filter design using a hamming window of order 80. The signals are next decimated from 120 Hz to 40 Hz to further focus on the spectral range of interest. Next, the spectra of each sensor’s signal is estimated by using sliding windows of 3 s and 2.75 s overlap with a 120-point discrete Fourier transform (DFT). The Welch power spectral density (PSD) estimation method with a Hamming window of 120 samples is used for PSD estimation, followed by a Gaussian-shaped moving average with a standard deviation of 1 Hz, to further smooth the spectra, sharpening the dominant frequencies and making them more distinguishable for the classifier. This results in 120 points of two-sided PSD with a spectral resolution of 0.33 Hz (40 Hz/120). The first 61 PSD values (corresponding to the DC component and one-sided spectrum) are used as the spectral feature vector of each sensor. The average feature vector calculated across all kinematic markers on a given extremity are then used as inputs to each of the classifiers described below.

##### B1: SVM classification

In algorithm B1, the 61-point one-sided average spectral features were directly provided to an SVM as feature vectors. We considered SVM models with both linear and radial basis function (RBF) kernels. A standard stratified 5-fold cross validation scheme was performed, by splitting the data into 5 nonoverlapping splits, using 4 splits for training and the left-out split for validation. The stratification ensured that each fold retained approximately equal proportions of the two class labels.

##### B2: XGBoost classification

In algorithm B2, the 61-point one-sided average spectral features were directly provided to XGBoost as feature vectors. XGBoost is also a widely recognized approach for classification tasks [26]. The XGBoost classifier operates by iteratively constructing an ensemble of decision trees and refining them based on a specified loss function. The procedure for loading the features was analogous to the SVM process, again using stratified 5-Fold cross-validation to ensure balanced representation across data splits. The classifier was configured to bypass label encoding, opting instead for the ‘logloss’ evaluation metric. This probability-centric metric enables the future extension of the proposed scheme for estimating probabilities of tremulous events, instead of a binary decision.

### 2.7. Performance Metrics

We compared the ability of six separate processing to recreate clinician ratings of tremor presence/absence based on *F*1 score, which is a harmonic mean of precision and recall [9]:

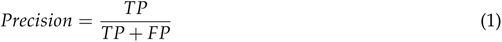

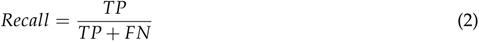

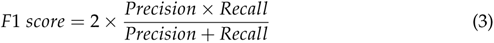

*TP* is a true positive that represents the total of successfully classified tremor-positive records, *FP* is a false positive that represents the total number of misclassified tremor-negative records, total misclassified class windows, and *FN* is a false negative that represents the total number of misclassified tremor-positive records.

We compared performance across classifiers using a confidence interval (CI) approach. For each of classifiers A1s, A2s, B1, and B2, we calculated the average (*µ*) and standard deviation (*σ*) of *F*1 scores observed across each of *n* = 5 folds during cross-validation. We then used the sample standard deviations to estimate 95% CIs as 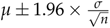. We applied similar analyses to secondary outcomes including Accuracy, Precision, Recall, and AUC. For classifiers A1r and A2r, we were only able to calculate point estimates of *F*1 and other outcomes as these classifiers were developed manually with access to the entire dataset.

Finally, we characterized the contributions of different frequency bands to classification with SHAP (SHapley Additive exPlanations) [27] plots. SHAP plots visually represent how much each feature contributes to the classification of each observation as one class or the other. This is analogous to the visual representation of factor loadings in familiar techniques like principal components analysis (PCA) but is adapted for nonlinear techniques like XGBoost.

## 3. Results

### 3.1. Characteristics of annotations

The most frequent clinical annotation was “present.” Clinicians used a wide range of qualitative labels for tremor size. A description of the mapping between raw clinician provided labels and dichotomized dataset labels is provided in Table 4. Annotations most frequently referred to the hands (37%), although annotations for all extremities were present. The frequencies of appearance of various body extremities are described in Table 5. Chi-squared tests identified significant differences in reporting frequencies between the arms and legs (P*≪*0.001) and between the arms and head/torso (P*≪*0.001).

**Table 4.** Mapping between clinician-provided labels and training labels in training data. The “Other” label aggregates annotations with fewer than 10 observations and annotations for which no indicator of size was provided (e.g., “RH tremor”).

| Raw Label | Label in Dataset |  |
| --- | --- | --- |
|  | Absent | Present |
| Absent | 1476 | 0 |
| Dystonia, dyskinesia, or other abnormal posture or movement | 68 | 0 |
| Present | 0 | 288 |
| Not much | 0 | 24 |
| Very slight or very trace | 0 | 10 |
| Slight or trace | 0 | 78 |
| Intermittent | 0 | 10 |
| Mild | 0 | 121 |
| Mild to moderate | 0 | 34 |
| Moderate | 0 | 55 |
| Moderate to severe | 0 | 17 |
| Significant | 0 | 32 |
| Severe | 0 | 17 |
| Other, or no indicator of size | 0 | 42 |

**Table 5.**
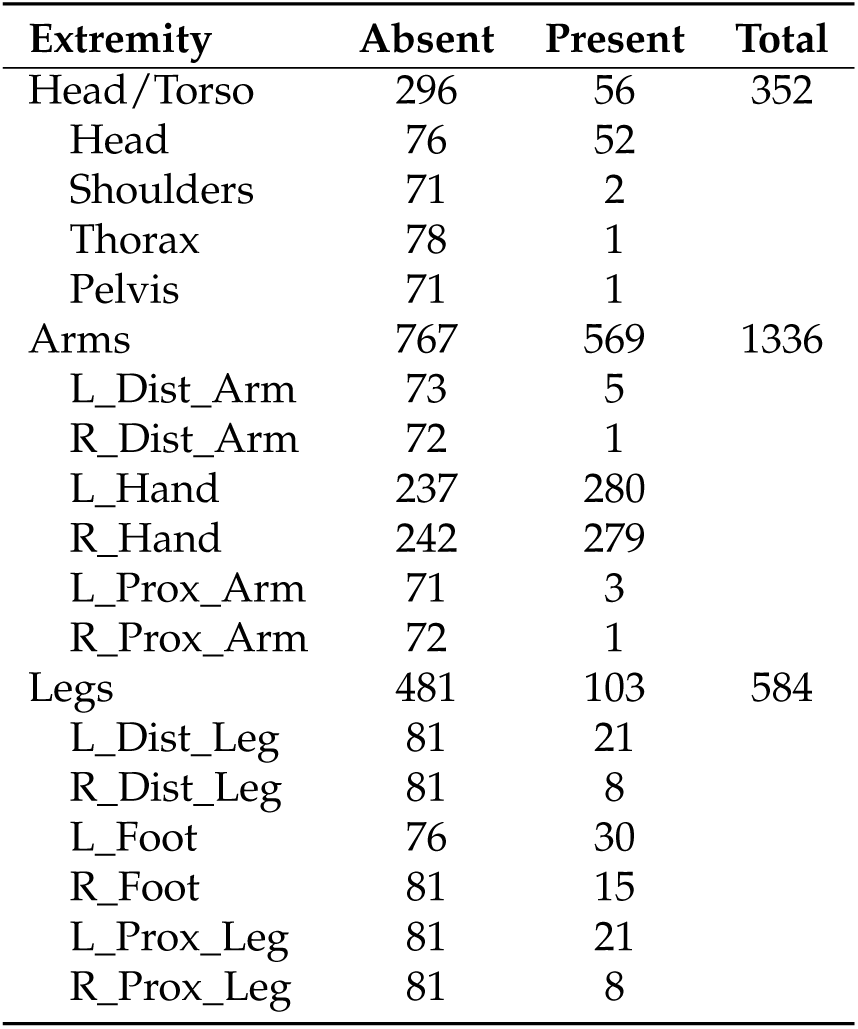
Frequency table of tremor annotations.

### 3.2. Model performance

Classification performance metrics for all models are reported in Table 6. The overall highest performance, as assessed with average *F*1 score across five cross-validation folds, was observed in algorithm A1s. However, all classifiers performed well. Across models, the average *F*1 score was 0.84 ± 0.02 (range [0.81 − 0.87]), and all *F*1 confidence intervals were overlapping. Figure 4 compares the performance of Algorithms B1 and B2 with operating point information superimposed for the other algorithms.

**Figure 4.**
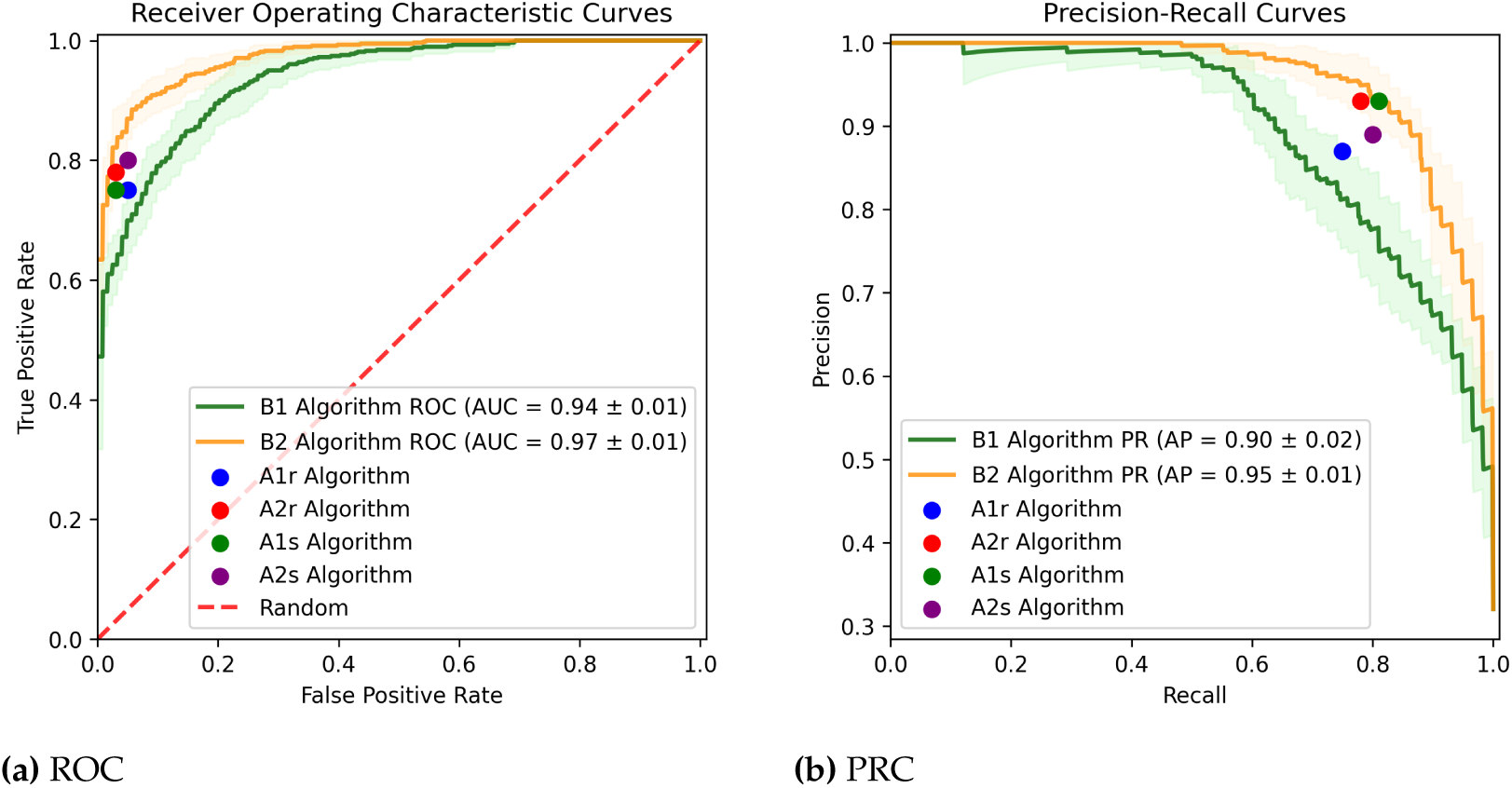
The average receiver operating characteristic (ROC) and precision-recall (PRC) curves for the SVM and XGBoost classifiers using spectral features of the spatial positions of the sensors. The shades correspond to *±*1 standard deviations of each curve across the five fold cross-validation. Refer to text for details.

**Table 6.**
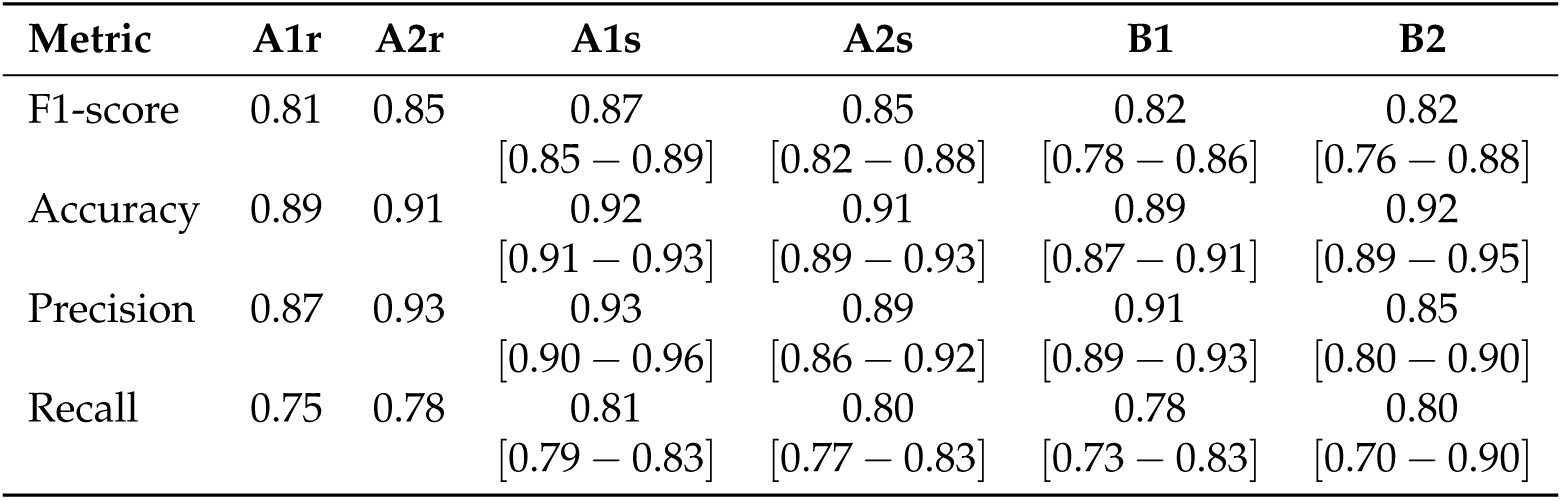
Algorithm performance comparison. Performance for hand-tuned algorithms A1r and A2r is reported across the entire dataset. Performance for other numerically optimized algorithms is reported across 5 separate data folds (Mean [95% CI]).

| Metric | A1r | A2r | A1s | A2s | B1 | B2 |
| --- | --- | --- | --- | --- | --- | --- |
| F1-score | 0.81 | 0.85 | 0.87<br>[0.85 – 0.89] | 0.85<br>[0.82 – 0.88] | 0.82<br>[0.78 – 0.86] | 0.82<br>[0.76 – 0.88] |
| Accuracy | 0.89 | 0.91 | 0.92<br>[0.91 – 0.93] | 0.91<br>[0.89 – 0.93] | 0.89<br>[0.87 – 0.91] | 0.92<br>[0.89 – 0.95] |
| Precision | 0.87 | 0.93 | 0.93<br>[0.90 – 0.96] | 0.89<br>[0.86 – 0.92] | 0.91<br>[0.89 – 0.93] | 0.85<br>[0.80 – 0.90] |
| Recall | 0.75 | 0.78 | 0.81<br>[0.79 – 0.83] | 0.80<br>[0.77 – 0.83] | 0.78<br>[0.73 – 0.83] | 0.80<br>[0.70 – 0.90] |

We further performed feature importance using SHAP values, for all spectral features used as inputs to algorithms B1 and B2. The SHAP plots are shown in Figure 5. Unlike typical SHAP plots that are oriented vertically, in Figure 5 the feature importance is shown on the y axis and the features - characteristic frequencies within the kinematic data - are shown on the x axis, which allows visualization of the SHAP plots as a kind of spectrum. We noted that high activation of features in the frequency range between 4.3 Hz and 7.0 Hz were identified as significant for identifying tremor presence vs. absence (red), while high activation of features in the frequency range between 0.7Hz and 1.3Hz were identifying tremor absence vs. presence (blue).

**Figure 5.**
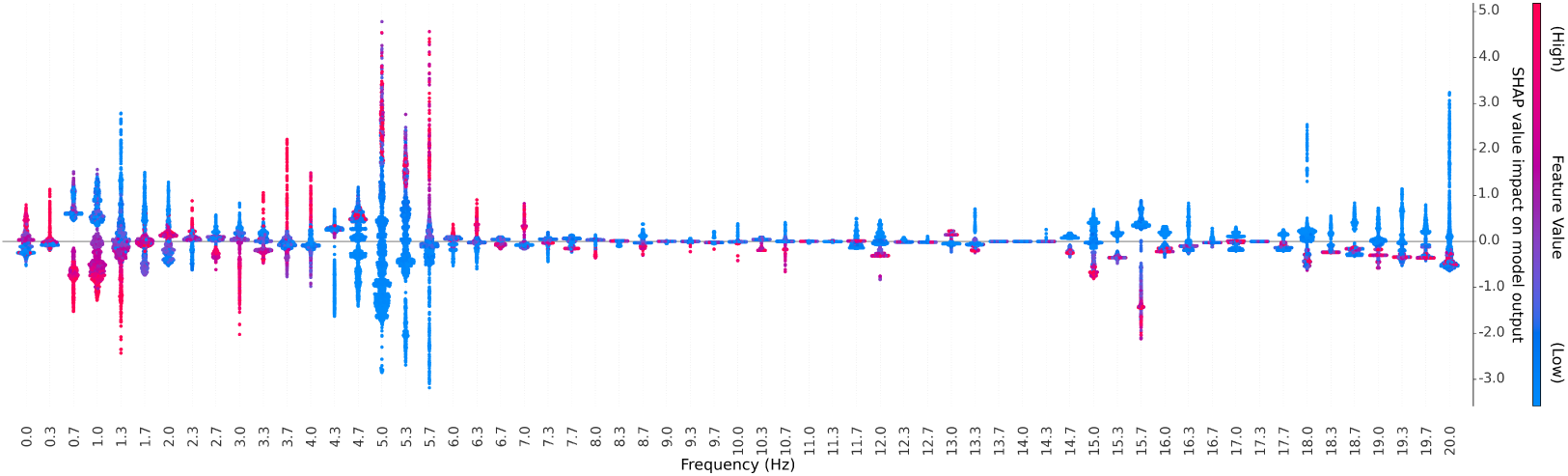
SHAP (SHapley Additive exPlanations) plot illustrating the contribution of each spectral feature across the Nyquist band, to the tremor prediction results. Each column on the plot represents a specific feature’s contribution to the prediction. Positive SHAP values drive the model’s output towards the tremor class, while negative values drive towards the non-tremor class. The color intensity indicates the magnitude of the feature value, with red denoting high values and blue indicating low values. Notice the significance of the frequency range between 4.3 Hz–7 Hz in identifying tremor. Frequencies below 3 Hz (corresponding to slow motions of the subject) are not informative for detecting tremor.

## 4. Discussion and Conclusion

The objective of this study was to assess the ability of several candidate processing pipelines to identify the presence or absence of tremor in kinematic data from movement disorders patients compared to expert ratings from movement disorders specialists. We found generally comparable performance across algorithms; the average *F*1 score was 0.84 0.02 (Mean SD; range 0.81 − 0.87), with all *F*1 confidence intervals overlapping. Notably, the highest performing algorithm (cross-validated *F*1 = 0.87) was Algorithms A1s, which was a version of the oldest algorithm in clinical use in our center that had been modified such that the manually-engineered features were used as inputs to a modern SVM with radial basis function kernels to accommodate linearly non-separable data.

These results suggest some points that may be generally useful in settings with site-specific, legacy clinical decision support systems. In particular, in our clinic’s implementation, the existing algorithms A1r and A2r lacked a clear separation between feature identification and classification steps. We anticipate that this may be the case in other centers with site-specific, legacy systems. Refactoring legacy code to separate these two separate steps may provide an important opportunity to introduce updated classifier architectures into these systems without discarding the rich domain knowledge that is embedded in the derivation of engineered features. In our case, although some of the engineered features (e.g., dominant frequency) could be trivially discovered by classifiers with generic spectral features (like B1 and B2), other features (e.g., symmetry of the dominant spectral peak) reflect clinical domain expertise that automated searches could miss given limited training data. We anticipate this as a common issue and recommend that centers utilizing legacy data processing routines refactor their algorithms to distinguish between feature extraction and classification to address this potential limitation and enhance algorithm performance.

Further, when we visualized the Receiver-Operator Characteristic curves (Figure 4), we found that the clinical algorithms A1r and A2r were tuned to penalize false positive rate at the expense of some sensitivity in clinical use. Because these algorithms were originally without hyperparameter tuning, this was not done intentionally on the part of the clinicians using these tools. Refactoring code could give clinicians the opportunity to tailor the balance of precision and recall to the clinical task at hand.

Analysis of SHAP plots revealed interesting information about the spectral composition of tremor. We noted that high activation of features in the frequency range between 4.3 Hz and 7.0 Hz were identified as significant for identifying tremor presence vs. absence (red), while high activation of features in the frequency range between 0.7Hz and 1.3Hz were identifying tremor absence vs. presence (blue) (Figure 5). This is consistent with literature using various sensing modalities that have described tremor [2] as producing frequency band activity around 5Hz, with voluntary movement producing lower frequency band activity below 3Hz. In particular, these results show that low frequency movement is not informative for detecting tremor; in fact, these frequencies have negative predictive value, suggesting that voluntary movements have the potential to be interpreted as false positives.

This was an informative result, as both clinical algorithms A1r and A2r were designed with features engineered to capture spectral information that was informative about tremor presence (presumably around 5Hz), but imposed no penalties on low frequency information that indicated that it was absent. This could be interpreted to mean that the original developers of these algorithms exhibited some cognitive confirmation or similar bias when designing the features to represent the aspects of the behavior they “knew about,” while neglecting equivalent kinematic information that was informative about the absence of tremor. The ability of modern ML to discover features may provide a unique opportunity to complement engineered features created with domain expertise.

### 4.1. Limitations

Our primary aim was to develop a generic tremor identification algorithm that could be used across extremities, behavioral tasks, and diagnoses. Although the resulting algorithm is almost certainly not optimal in all settings, this approach generally aligns with clinical best practices and represents an important first step in the development of a comprehensive clinical decision support tool for tremor. However, this necessarily comes with the limitation that this tool may not be appropriate for all tremor identification tasks.

Further, our tremor assessment approach does not use scripted voluntary movements and weight application in order to isolate mechanical, volitional, and pathophysiological causes of tremor [28].

### 4.2. Unique contributions

Our method assesses tremor across the body, a unique capability. In a recent machine learning review on tremor applications, only 14% (5/37) explored body parts beyond hands or distal arms [1]. This instrumentation is certainly convenient and almost certainly sufficient for tremor characterization with frequency [2] or amplitude [29,30]. However, we know that signal processing approaches like correlation across body regions provide additional diagnostic insight for discriminating, for example, parkinsonian from orthostatic tremors [28]. With full body data, end-to-end machine learning approaches (e.g., [9]) have significant potential to discover these and other features automatically. Other more subtle tremor features like distractibility [28] seem more likely to be characterized in full body data. Further, our testing approach imposes few, if any constraints on the participants’ natural movements. This complicates data analysis compared to methods that confine movements to a single plane, (e.g., [13]) but may improve external validity.

### 4.3. Conclusion

Here, we sought to assess the ability of several candidate processing pipelines to identify the presence or absence of tremor in kinematic data from movement disorders patients compared to expert ratings from movement disorders specialists. We found that many solutions offered acceptable performance. The best individual-performing algorithm was a modernization of one of the oldest algorithms in constant clinical use in our center. In general, updating legacy clinical decision support systems to incorporate modern machine learning classifiers may result in better performing tools and associated decreases in provider time and improved outcomes.

## Institutional Review Board Statement

The study was conducted in accordance with the Declaration of Helsinki, and approved by the Institutional Review Board of Emory University under IRB protocol 00002688 approved June 2, 2021.

## Informed Consent Statement

Patient consent was waived due to the retrospective nature of this study.

## Data Availability Statement

Data are available upon reasonable request from the corresponding author.

## Disclaimer/Publisher’s Note

The statements, opinions and data contained in all publications are solely those of the individual author(s) and contributor(s) and not of MDPI and/or the editor(s). MDPI and/or the editor(s) disclaim responsibility for any injury to people or property resulting from any ideas, methods, instructions or products referred to in the content.

## Data Availability

All data produced in the present study are available upon reasonable request to the authors

## Abbreviations

CPT: Current Procedural Terminology
ET: Essential tremor
FTM: Fahn-Tolosa-Marin Tremor Rating Scale
MDS-UPDRS-III: Movement Disorder Society-Unified Parkinson’s Disease Rating Scale Part III
PD: Parkinson’s disease
SHAP: SHapley Additive exPlanations
SVM: Support vector machine
TETRAS: The Essential Tremor Rating Scale

## Appendix A Comparison of tremor features identified by algorithms A1r and A2r

We performed some additional analyses to compare the tremor features identified by clinical algorithms A1r and A2r. We compared tremor frequencies identified by clinical algorithms A1r and A2r using a Bland-Altman approach [31]. Because each of the clinical algorithms produced an estimate of tremor amplitude whether or not a tremor was detected, we compared the ranges of amplitudes obtained when tremor was present or absent according to ground-truth labels with two-sample Kolmogorov-Smirnov tests.

Overall, algorithms A1r and A2r identified very similar central tremor frequency estimates, with average values 4.8 ± 1.0 Hz and 4.7 ± 0.6 Hz, respectively. Bland-Altman analysis between the results of the two algorithms identified a bias of 0.1 Hz between the two algorithms, with 95% limits of agreement ( −9.0, 0.7) Hz (Figure A.1).

With both algorithms, the range of identified amplitudes for which tremor was rated present according to expert labels had some overlap with the range of amplitudes for which tremor was rated absent. This suggests that a simple amplitude-based threshold would be insufficient to discriminate tremor presence using either approach. With algorithm A1r, the average amplitude when tremor was present was [102.2 13.40, 1.9 944.7] mm/s ([Mean ± SD, range]) compared to [26.4 35.1, 0.4 199.0] mm/s when tremor was absent. With algorithm A2r, the average amplitude when tremor was present was [3.07 3.00, 0.3 24.3] mm compared to [0.13 0.12, 0.01 0.59] mm when tremor was absent. The cumulative densities identified by both algorithms showed separation between cases labeled as tremor absent and present and highly significant (P 0.001) two-sample Kolmogorov-Smirnov tests. Visual inspection of cumulative amplitude distributions (Figure A.2) for the two algorithms suggested that A2r provided better separation, although this could not be compared directly due to the different units used by the algorithms.

**Figure A.1.**
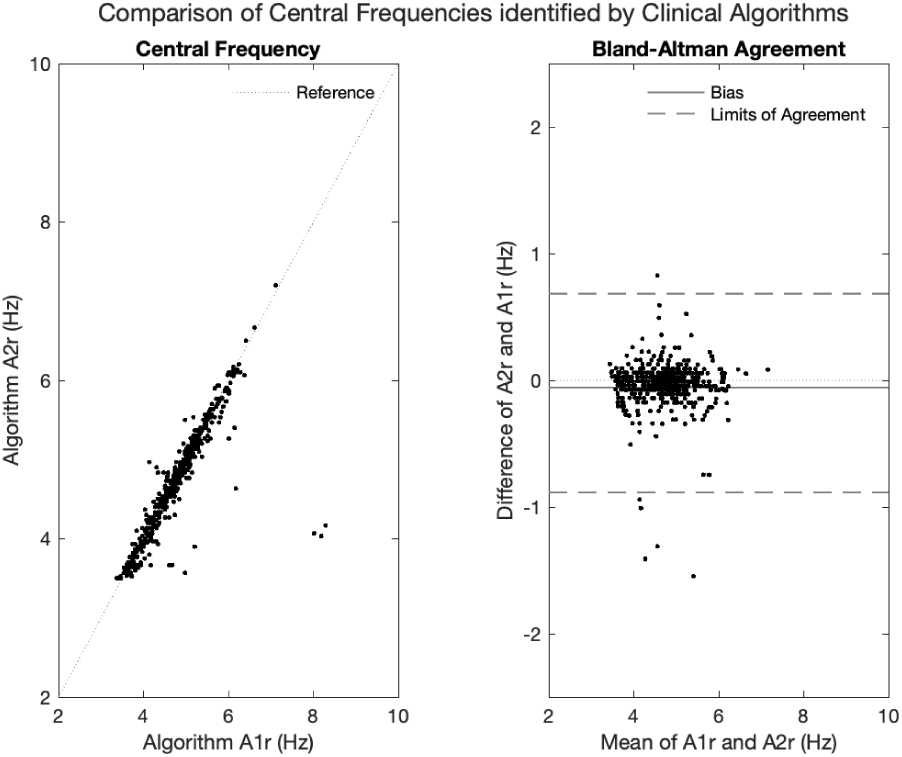
Comparison of tremor frequencies identified by clinical algorithms A1r and A2r.

**Figure A.2.**
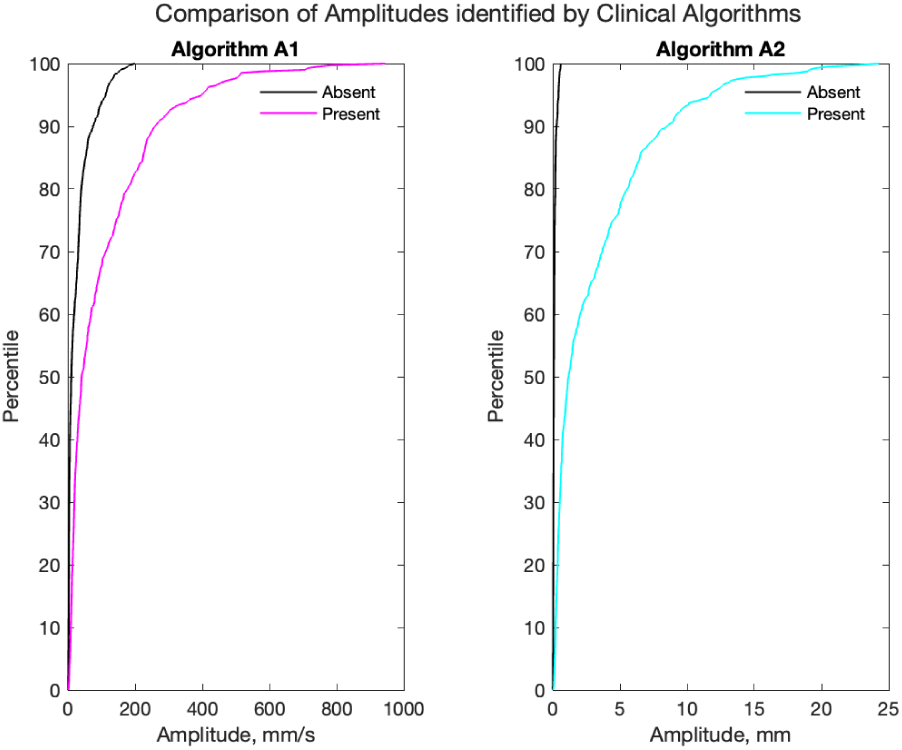
Comparison of tremor amplitudes identified by clinical algorithms A1r and A2r, stratified by ground-truth labels of tremor presence or absence.

## Appendix B Additional dataset details

This section provides some additional details on the data format and coding scheme. During each individual behavioral test, the laboratory 3D motion capture system records the instantaneous position of all kinematic markers on the body (typically 60) and exports these data to a standard *\*.trc* tabular data format with some minimal header information. A typical *.trc* file for a 30-second recording at 120 Hz comprises 3600 rows (30 seconds × 120 Hz) and 180 columns (60 markers × 3 axes) of kinematic data. Because each file includes data from markers on different extremities, for which tremor may be absent or present on a give trial, the columns of the *\*.trc* file corresponding to markers on each extremity must be separated prior to analysis. Our clinical data processing pipeline maintains the mappings between kinematic markers and extremities in an *\*.xml* file (markers.xml). To avoid the burden of parsing these files, the data supplied with the paper are provided in two different formats. Each deidentified *.trc* file is provided as originally exported, as well as divided into separate *.csv* files for each body extremity in the accompanying dataset. These files are compatible with standard Python, R, Matlab, or similar software libraries. Summaries of the contents of example files are provided in Table B.1. Descriptions of kinematic marker locations are provided in Tables B.3 through B.5.

Information about the testing condition used during each recording is provided as part of the individual file names, using the nomenclature provided in Table B.2. For example, the file *data/HH/std-arms-extended1-TP.trc* designates participant HH standing with arms extended forward along the X axis of the laboratory 1. In pointing and spiral movement tasks, the suffix *right* or *left*, or *1* or *2*, are appended to the filename denoting the extremity involved. In some cases, extra motor or cognitive tasks were introduced to intensify tremor provocation, with supplementary information appended to the base task codes.

**Table B.1.**
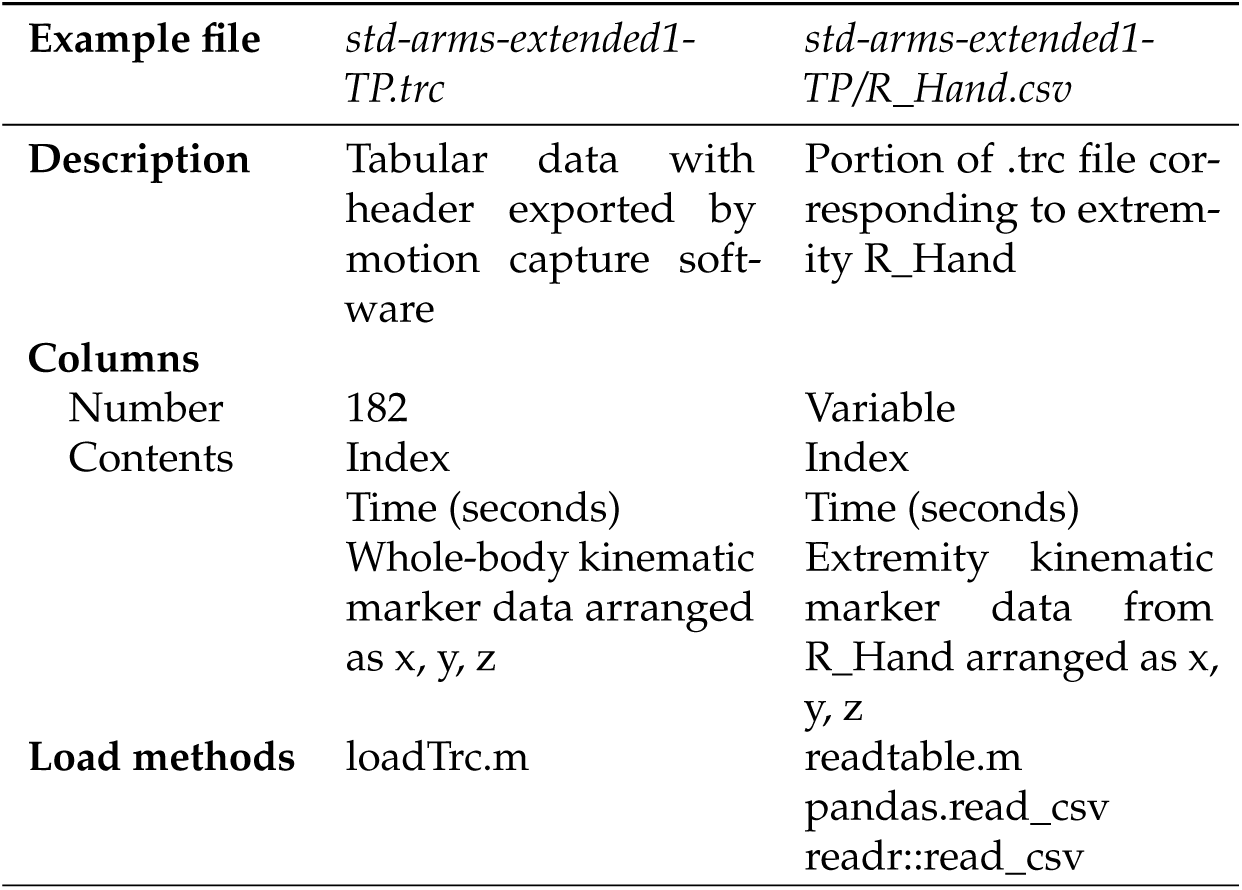
Example file descriptions and load methods. In some cases .trc files contain additional columns with derived variables that should be ignored.

| Example file | *std-arms-extended1-TP.trc* | *std-arms-extended1-TP/R_Hand.csv* |
| --- | --- | --- |
| **Description** | Tabular data with header exported by motion capture software | Portion of .trc file corresponding to extremity R_Hand |
| **Columns** |  |  |
| Number | 182 | Variable |
| Contents | Index<br>Time (seconds)<br>Whole-body kinematic marker data arranged as x, y, z | Index<br>Time (seconds)<br>Extremity kinematic marker data from R_Hand arranged as x, y, z |
| **Load methods** | loadTrc.m | readtable.m<br>pandas.read_csv<br>readr::read_csv |

**Table B.2.** Nomenclature for behavioral tasks employed in testing.

| Code | Task |
| --- | --- |
| sit-rest | Seated with arms at sides |
| sit-arms-extended | Seated, with arms extended anteriorly and parallel to the floor |
| sit-UEopp | Seated, with arms in a “T” pose parallel to the ground with fingers of each hand opposed |
| sit-point | Seated, performing a finger-to-nose pointing task with the indicated extremity (*right/1* or *left/2*) |
| sit-spiral | Seated, performing a spiral movement with the indicated extremity (*right/1* or *left/2*) |
| std-rest | Standing with arms at sides |
| std-arms-extended | Standing, with arms extended out parallel to the ground |
| std-UEopp | Standing, with arms in a “T” pose parallel to the ground with fingers of each hand opposed |
| walk-thru | Comfortable walking from one end to the other of the motion capture space |
| TUG | Sequential “timed up and go” walking tasks |

**Table B.3.** Kinematic marker descriptions for markers on the trunk. Markers that appear on both sides of the body are listed for the right side only and are coded beginning with “R.” Replacing this character with “L” will designate the corresponding marker on the left side of the body.

| Extremity | Marker Code | Description |
| --- | --- | --- |
| Head | Front_Head | Center of forehead, on cap |
|  | JAW | Mental protuberance |
|  | Jaw | Mental protuberance |
|  | RBHD | Right back head, on cap |
|  | RFHD | Right front head, on cap |
|  | Rear_Head | Rear of head, on cap |
|  | TopHead | Top of head |
|  | Top_Head | Top of head |
| Shoulders | C7 | Seventh cervical vertebra |
|  | RBAK | Right scapula (asymmetry marker) |
|  | RSHO | Right acromioclavicular joint |
|  | R_Shoulder | Right acromion process |
|  | STRN | Xiphoid process |
| Pelvis | LASI | Left anterior superior iliac spine |
|  | RASI | Right anterior superior iliac spine |
|  | RIC | Right iliac crest |
|  | RPSI | Right posterior superior iliac spine |
|  | R_ASIS | Right anterior superior iliac spine |
|  | V_Sacral | Sacrum |
| Thorax | CLAV | Clavicular notch |
|  | R_Clavicle | Right clavicle |
|  | R_Scap_Inf | Right scapula inferior angle |
|  | R_Scapula | Right supraspinous fossa |
|  | T10 | 10th thoracic vertebra |

**Table B.4.**
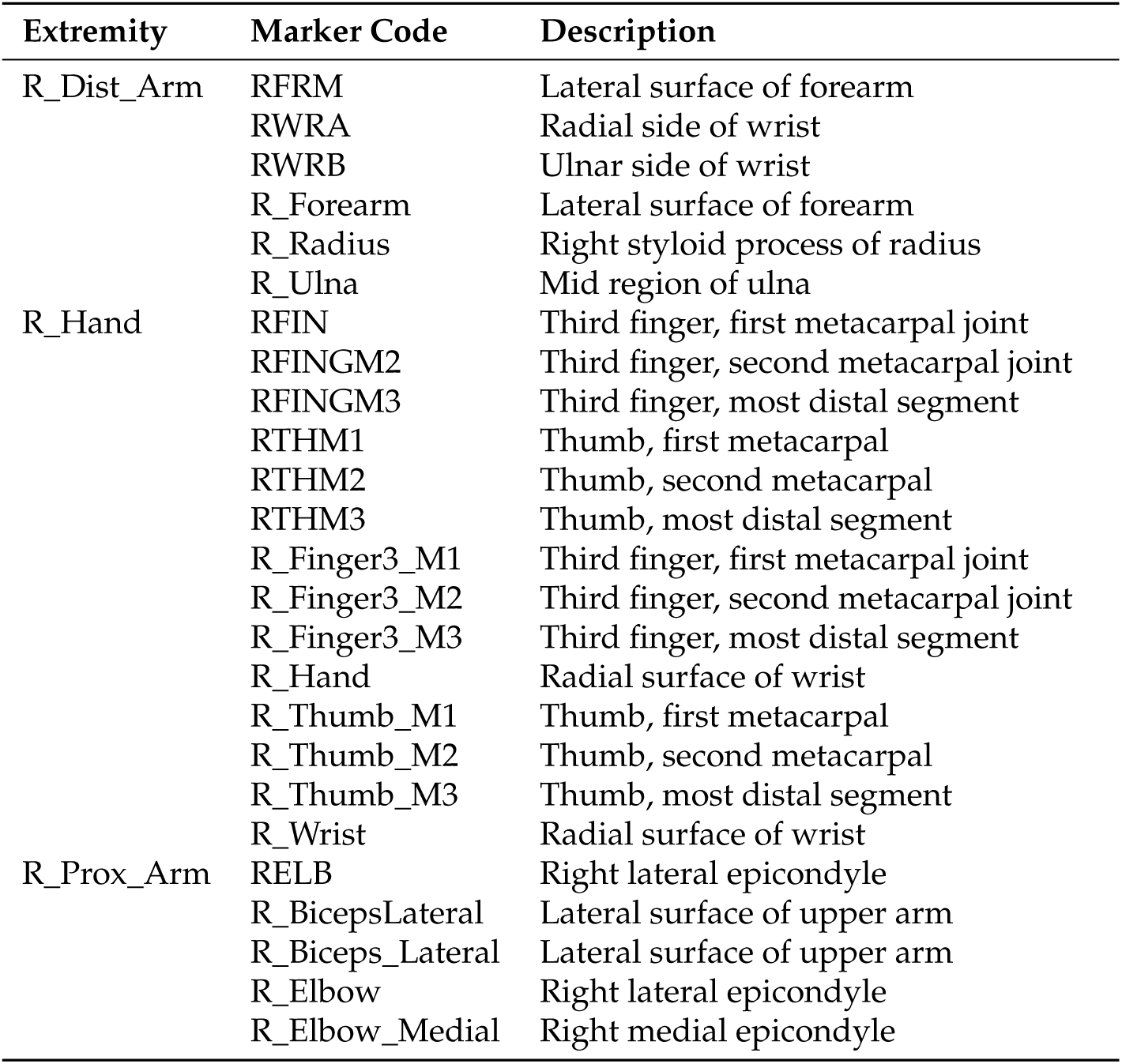
Kinematic marker descriptions for markers on the arms. Markers that appear on both sides of the body are listed for the right side only and are coded beginning with “R.” Replacing this character with “L” will designate the corresponding marker on the left side of the body.

**Table B.5.**
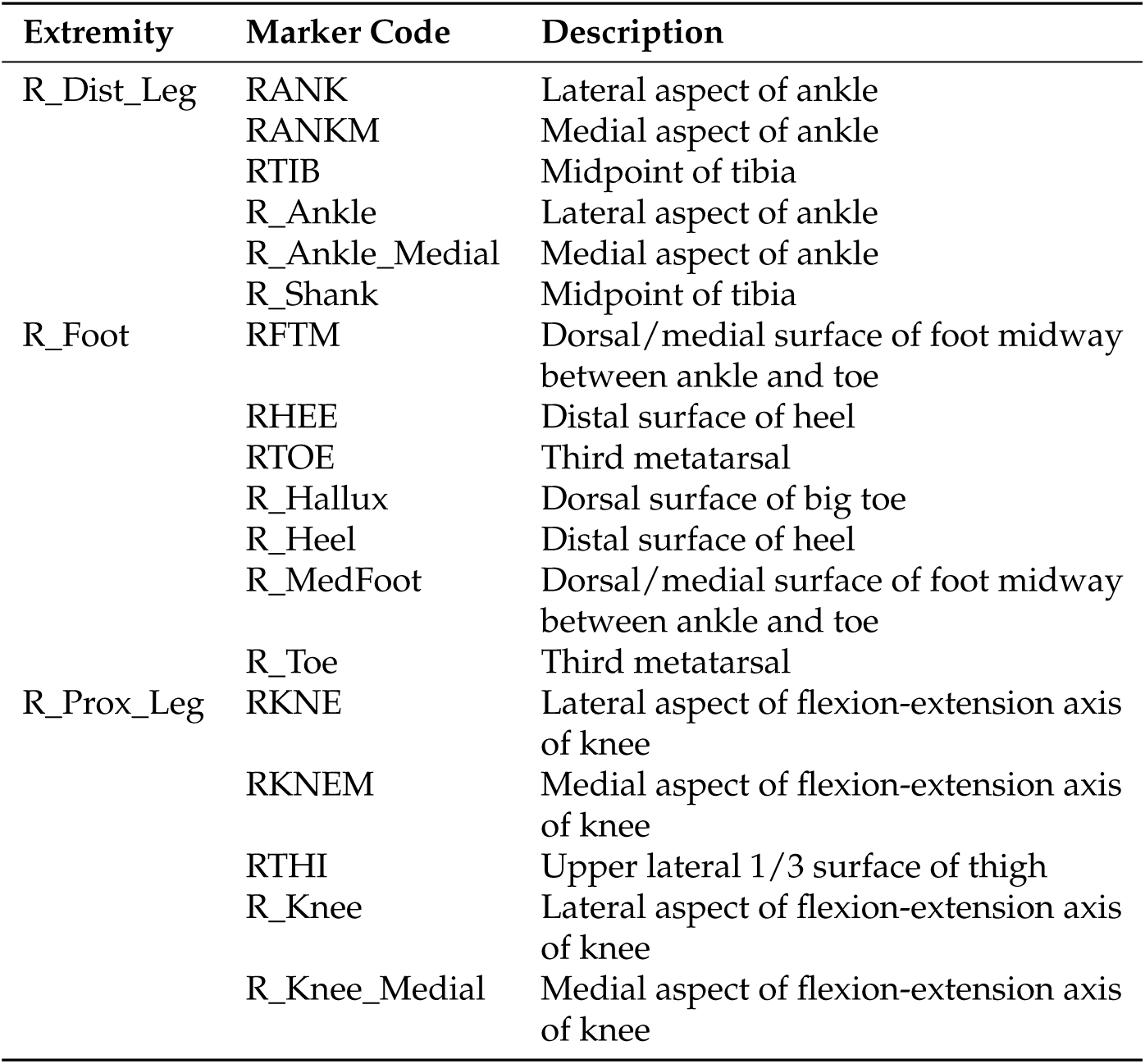
Kinematic marker descriptions for markers on the legs. Markers that appear on both sides of the body are listed for the right side only and are coded beginning with “R.” Replacing this character with “L” will designate the corresponding marker on the left side of the body.

